# Dynamic Analysis on Simultaneous iEEG-MEG Data via Hidden Markov Model

**DOI:** 10.1101/2020.07.22.20159566

**Authors:** Siqi Zhang, Chunyan Cao, Andrew Quinn, Umesh Vivekananda, Shikun Zhan, Wei Liu, Bomin Sun, Mark Woolrich, Qing Lu, Vladimir Litvak

## Abstract

**Background:** Intracranial electroencephalography (iEEG) recordings are used for clinical evaluation prior to surgical resection of the focus of epileptic seizures and also provide a window into normal brain function. While these recordings afford detailed information about local brain activity, putting this activity in context and comparing results across patients is challenging. Non-invasive whole-brain Magnetoencephalography (MEG) could help translate iEEG in the context of overall brain activity, and thereby aid group analysis and interpretation.

**Methods:** Simultaneous MEG-iEEG recordings were performed at rest on 11 patients with epilepsy. Pre-processed MEG sensor data was projected to source space. The time delay embedded hidden Markov model (HMM) technique was applied to find recurrent sub-second patterns of network activity in a completely data-driven way. To relate MEG and iEEG results, correlations were computed between HMM state time courses and iEEG power envelopes in equally spaced frequency bins and presented as correlation spectra for the respective states and iEEG channels.

**Results:** Five HMM states were inferred from MEG. Two of them corresponded to the left and right temporal activations and had a spectral signature primarily in the theta/alpha frequency band. The majority of iEEG contacts were also located in left and right temporal areas and the theta/alpha power of the local field potentials (LFP) recorded from these contacts correlated with the time course of the HMM state corresponding to the temporal lobe of the respective hemisphere.

**Discussion:** Our findings are consistent with the fact that most subjects were diagnosed with temporal epilepsy and implanted with temporal electrodes. As the placement of electrodes between patients was inconsistent, their modulation by HMM states could help group the contacts into functional clusters. This is the first time that HMM was applied to simultaneously recorded iEEG-MEG and our pipeline could be used in future similar studies.

## INTRODUCTION

In patients with focal refractory epilepsy, surgical resection of the epileptic focus may be a therapeutic option. Intracranial electroencephalography (iEEG) is an invasive procedure involving implantation of electrodes into the brain to guide surgical planning (Assi et al., 2019). Despite possessing excellent temporal resolution, iEEG is associated with sparse spatial sampling due to the limited number of electrodes implanted (Velmurugan et al., 2019). Magnetoencephalography (MEG) has been an increasingly utilised non-invasive method in surgical pre-evaluation of focal epilepsy, providing high temporal and spatial resolution and the whole-brain context (Gavaret et al., 2016) to abnormal epileptic activity.

Combined MEG and iEEG recordings performed at different timepoints showed that MEG could noninvasively identify regional interictal networks (Stefan and Trinka, 2017). iEEG implantation guided by MEG findings increases the likelihood of successful resection (Murakami et al., 2016). Several studies used acquisitions of both iEEG and MEG to explore the accuracy of MEG for localizing the epileptic focus (Kim et al., 2016), the contribution of MEG for identifying iEEG implantation sites (Agirre-Arrizubieta et al., 2014) and the correspondence between MEG and iEEG in the presumed epilepsy focus (Grova et al., 2016). When comparing iEEG and EEG/MEG, MEG measurements were shown to be able to extract a significant proportion of epilepsy networks visible in iEEG (Malinowska et al., 2014). Comparing localisation results for epileptic spikes and oscillations between the two modalities showed better concordance for spikes (Jmail et al., 2016). In addition to the clinical application in epilepsy, other studies used this multimodal approach to investigate spatiotemporal profiles of word processing (McDonald et al., 2010) or the relationships of fast- and slow-timescale brain oscillatory dynamics (Zhigalov et al., 2015).

In most of these studies, the recordings were performed separately for each modality. Thus, the relationship between neural oscillations recorded at various scales remains poorly understood partly due to the technical difficulty associated with acquiring simultaneous multimodal brain recordings (Dalal et al., 2009). Such recordings make it possible to explore the consistency between modalities, when the exact same brain states are assessed by both (He et al., 2019). Simultaneous acquisition of non-invasive MEG and iEEG data is also extremely valuable for validation of MEG methods

The sensitivity of whole-head MEG versus iEEG in the detection and localization of epileptic spikes was simultaneously assessed and validated by (Santiuste et al., 2008). Another study evaluated the relationship between the amplitude recorded from iEEG electrodes in the lateral temporal region, and their distance from the MEG-modelled spikes (Kakisaka et al., 2012). Recently it was shown using simultaneous recordings that both MEG and iEEG could detect epileptogenic activity from deep sources such as amygdala and hippocampus (Pizzo et al., 2019).

The brain in the resting state is not quiescent. Previous electrophysiological studies have revealed that resting state networks are underpinned by rich spatiotemporal dynamics (Brookes et al., 2014; O’Neill et al., 2018), which could be characterized using time-varying measures of interactions (Chang et al., 2013; Zhang et al., 2018). Analyses using sliding time-window approaches on both resting data (de Pasquale et al., 2012) and task data (O’Neill et al., 2017) must solve the problem of determining the window length. One method to define resting state networks without pre-specification of the sliding window length is Hidden Markov model (HMM). This method was shown to be able to infer a number of discrete brain states that recur at different points in time on a sub-second temporal scale (Baker et al., 2014). A novel HMM method, referred to as time-delay embedded HMM (TDE-HMM), could not only identify large-scale phase-coupled network dynamics, but also characterize spatial patterns of oscillatory power and coherence in specific frequency bands (Vidaurre et al., 2018).

Here we apply a group-level TDE-HMM analysis to MEG and iEEG data recorded simultaneously in epilepsy patients at rest. Our aims were to show as a proof of principle that this analysis is possible in patients with possibly abnormal and inconsistent functional anatomy and to examine invasive correlates of non-invasively identified network states. We found large-scale resting-state networks consistent with those previously described for healthy subjects. While each state had subject-specific temporal characteristics, the spatial and spectral features of all states were well-defined at the group level. Network states identified with TDE-HMM in MEG data correlated with spectral modulations in iEEG data.

## METHODS

### Participants

Simultaneous MEG-iEEG recordings were performed on 11 patients with intractable epilepsy undergoing pre-surgery evaluations. The patients were recruited from the Department of Neurosurgery, affiliated Ruijin Hospital, Shanghai Jiao Tong University School of Medicine. Intracranial electrodes were implanted for pre-resection seizure localization guided strictly by clinical indications.

### Ethics Statement

The study was approved by the local ethics committee of Ruijin hospital, Shanghai Jiaotong University School of Medicine and in accordance with The Code of Ethics of the World Medical Association (Declaration of Helsinki) for experiments involving humans. Every patient was informed about the aim and the scope of the study and gave written informed consent.

### Data acquisition

Implantation of the depth electrodes (SDE-08: S8 and S16, Beijing Sinovation Medical Technology CO., LTD, Beijing, China) was performed under general anaesthesia. Location and number of iEEG electrodes implanted varied between patients depending on presumed epileptogenic focus. Table 1 summarizes the patients’ clinical and iEEG characteristics. Resting MEG recordings were carried out using the Elekta Neuromag Vector View 306 channel System in a magnetically shielded chamber. The EEG system integrated with the MEG was used for the iEEG recordings. The patients were instructed to rest with eyes closed.

**Table 1.** Clinical and iEEG characteristics of epilepsy patients

| Case | Age<br>(years) | Gender<br>(M/F) | Epilepsy duration<br>( Months ) | Locations of electrodes | #Contacts<br>#Electrodes |
| --- | --- | --- | --- | --- | --- |
| 1 | 24 | F | 2 | r temp, l temp, r parietal, l parietal | 32/4 |
| 2 | 55 | F | 15 | l hippo, l temp, l parietal, r hippo, r temp | 48/6 |
| 3 | 47 | F | 10 | r hippo, r temp, r parietal | 32/4 |
| 4 | 14 | M | 6 | r front, r parietal, r insular | 32/4 |
| 5 | 33 | F | 14 | l temp, l parietal, hippo | 32/4 |
| 6 | 24 | F | 6 | r temp, r parietal | 24/3 |
| 7 | 26 | F | 2 | l temp, l insular, l occip, r temp | 48/5 |
| 8 | 19 | F | 10 | l hippo, l parietal, l occip, r hippo | 48/6 |
| 9 | 33 | M | 9 | l hippo, l temp, l parietal, l front | 64/6 |
| 10 | 32 | F | 14 | l hippo, l temp, l front | 48/4 |
| 11 | 27 | F | 10 | l hippo, l temp, l lingual, l occip, l cicu, r hippo | 64/8 |

### Code and data availability statement

The code used for the following analysis presented here is available at https://github.com/SiqiZhang0106/Dynamic-HMM-Analysis-on-Simultaneous-iEEG-MEG. Data sharing is subject to ethics restrictions and therefore the data will be shared on request addressed to Dr. Chunyan Cao and subject to data sharing agreement.

### Data Analysis

Anatomical data were processed with the Lead-DBS toolbox (http://www.lead-dbs.org/) (Horn and Kuhn, 2015) to reconstruct the contact locations. iEEG contact locations were obtained by fusing a post-operative CT scan with a pre-operative T1 structural MRI scan and manually fitting electrode models to the artefacts seen in the CT. The electrode locations were then transformed to standard stereotactic space used in Lead DBS (MNI 2009b NLIN asymmetric template).

The MEG data were de-noised using Maxfilter™ software implementing the temporal extension of the signal space separation method (tSSS) (Taulu and Hari, 2009). Interictal spikes were identified by a trained clinician and segments of ±5sec around spikes were excluded from analysis. The subsequent analyses were performed using the Oxford centre for Human Brain Activity (OHBA) Software Library (OSL) (https://ohba-analysis.github.io/osl-docs/) (Quinn et al., 2018). This builds upon Fieldtrip (http://www.fieldtriptoolbox.org/) (Oostenveld et al., 2011) and SPM (http://www.fil.ion.ucl.ac.uk/spm/) toolboxes (Litvak et al., 2011). Structural MRI and the MEG data were co-registered by RHINO (Registration of Headshapes Including Nose in OSL). The MEG data were then downsampled to 250 Hz and filtered to the frequency band from 1 to 45 Hz. Time segments containing artifacts were detected using the generalized extreme studentized deviate method (Rosner, 1983) to reject outliers in the standard deviation of the signal computed across all sensors. Subsequently, temporal independent component analysis (ICA) produced independent components that were visually checked to remove artifacts related to breath, heart beats, movement and muscle activity (4.3 ± 2.1 components (mean ± SD) were removed from each data session). A Linearly Constrained Minimum Variance (LCMV) vector beamformer was applied on the pre-processed sensor data to project them onto an 8mm grid in source space (Van Veen et al., 1997; Woolrich et al., 2011). Parcel-wise time series with 39 regions covering the entire cortex were estimated by taking the first component of a weighted PCA across voxels within each parcel (Quinn et al., 2018). A multivariate symmetric orthogonalization was then adopted to attenuate the spatial leakage effects (Colclough et al., 2015) including those caused by ghost interactions (Palva et al., 2018).

### HMM Inference

Time Delay Embedded HMM technique (TDE-HMM) finds recurrent patterns of network (or HMM state) activity in a completely data-driven way. Each state could be characterised by specific patterns of power and phase-coupling connectivity (Quinn et al., 2018; Vidaurre et al., 2018). The TDE-HMM is inferred from the source-space MEG data using the HMM-MAR toolbox (https://github.com/OHBA-analysis/HMM-MAR). The time course of each parcel was embedded with a time delay using L lags. L was set to be 15 following Quinn et al. (2018) with lags between -7 and 7 time steps. As we had previously downsampled the data to 250 Hz, L of 15 lags corresponded to 30 ms lags in both directions and resulted for each subject in an extended data matrix of (L lags * N nodes) * S timesamples. The first dimension of the matrix was reduced from 15×N to 4×N by principal component analysis. The HMM-MAR uses stochastic inference, the batch size was set to use 15 continuous data segments at each iteration, this is based on taking subsets or batches of subjects at each iteration instead of the entire data set. And maximum number of variational inference cycles was set to 500. The whole HMM inference was run 10 times to ensure the stability of results and the best performance with lowest free energy was accepted here. The state observation models were characterized by multivariate auto- and cross-covariance matrices and a time series of posterior probabilities was inferred to represent the occurrence probability of a state at a time point. A Viterbi Path (Bishop, 2006) represented the mutually exclusive state allocations.

For each inferred state, corresponding probability time course was computed and the state-specific MEG power spectrum was estimated in the range of 1-30Hz. To aid visualization, spectral modes were then generated by computing a Non-Negative Matrix Factorisation (NNMF) across the spectral estimates (Quinn et al., 2018).

### Establishing the relation to iEEG data

To interrogate the simultaneously acquired intracranial data, a ‘correlation spectrum’ was computed for time series of each iEEG bipolar channel and each HMM-derived state. This computation was done as follows. First, time-frequency decomposition was calculated for each iEEG bipolar channel using the same time resolution as for HMM pre-processing of MEG to achieve sample-by-sample correspondence between HMM state time courses and iEEG power time courses. After that, the correlations between each HMM-derived state probability time course and each frequency bin of the iEEG time-frequency matrix were computed. Finally, the correlation coefficients were plotted as a function of frequency, resulting in a separate correlation spectrum for each state and bipolar channel.

In addition, to group the iEEG contacts into HMM-related clusters, we determined the spatial relationship of each iEEG contact and each MEG-inferred HMM state activation. As mentioned above, HMM inference and output spectral maps were all based on 39 brain parcels. MNI coordinates of voxels within each parcel could be acquired from the template. Each state corresponded to several activated parcels (with the top 10% of activations). Therefore, the Euclidean distance between each contact and voxels within each state’s parcels could be calculated and averaged, resulting in a distance between the contact and the state. If a contact had a smaller distance with a particular state compared with the other states, it would be assigned to this state.

### Comparison to HMM derived from a healthy subject dataset

Although epochs of abnormal interictal activity were removed, slow wave activity associated with epilepsy could possibly still affect inference of the MEG-derived HMM state models. Thus, the five HMM states derived from the patient MEG data were validated by using an HMM inferred on a large number of healthy subjects acquired as part of a different project at OHBA. This healthy dataset was also recorded in a Neuromag 306 MEG system, and was pre-processed and the TDE-HMM was inferred in the same way as for our patient analysis. We then sought to estimate the state-specific spectral group activation maps in the patient MEG that corresponded to the HMM states inferred on the healthy cohort. First, we extracted the state-specific group observation models from the HMM inferred on the healthy dataset; and then, holding them fixed, estimated the state time-courses (i.e. the occurrence of the states) in the epilepsy patient dataset. Finally, using these state time courses, the state-specific spectral activation group maps were estimated for the patient MEG data.

The entire workflow is illustrated in Figure1.

**Figure 1.**
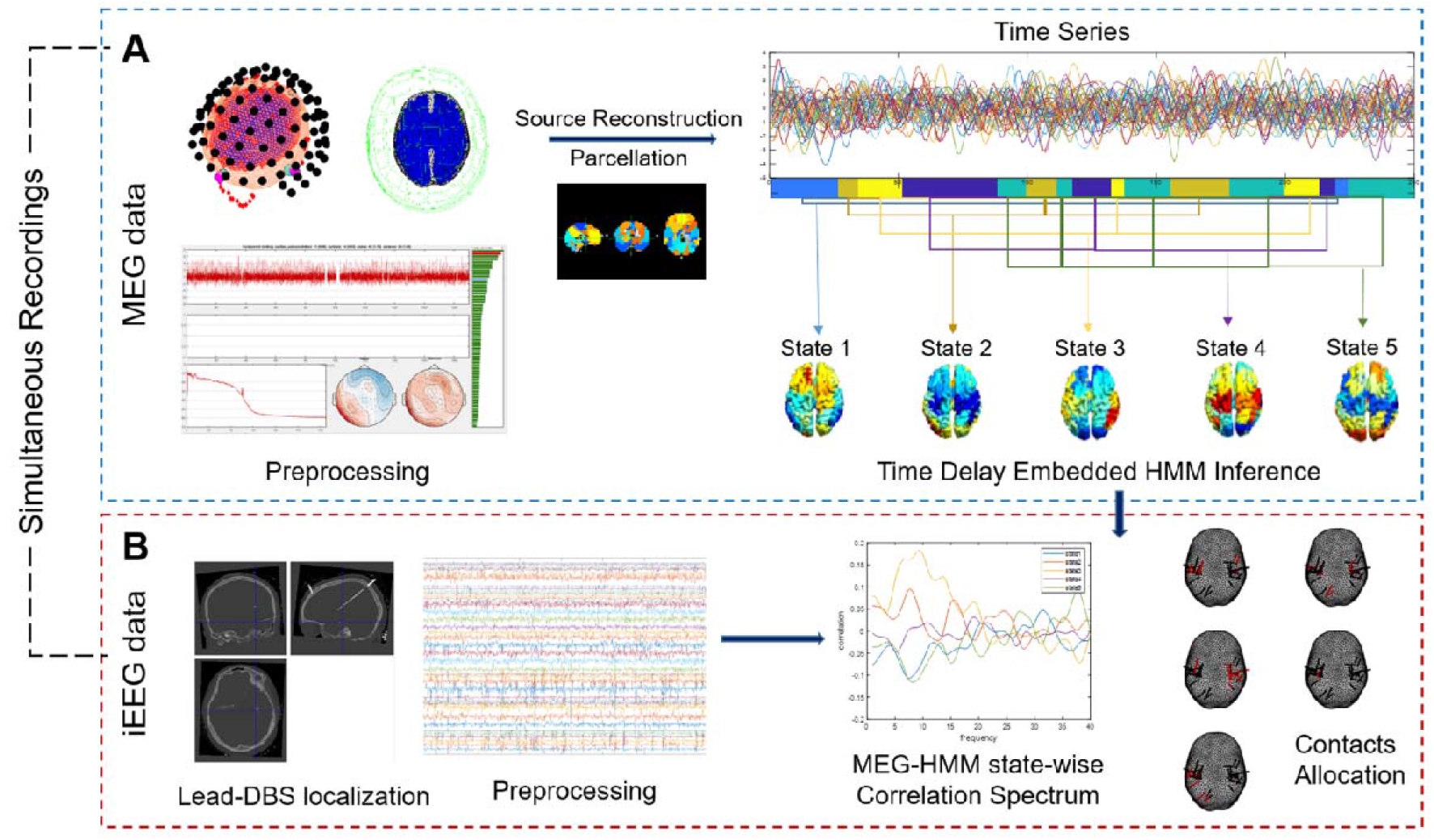
The entire workflow including HMM inference on MEG data and relating the resulting states to iEEG data A. MEG sensor-data were pre-processed and source reconstructed. Then concatenated time series of all subjects were used to infer the time delay embedded HMM states. B. Simultaneous iEEG data were pre-processed and subjected to the same time-frequency decomposition as MEG. The iEEG power time courses were correlated with MEG-HMM probability matrix (see Fig 3A for details). iEEG leads were localised and bipolar channels were assigned to states according to their spatial correspondence (see Fig 3B for details).

## RESULTS

After extracting and concatenating MEG data of 11 subjects, we identified 5 HMM states using TDE-HMM. Prior to that we tested a range of values for the number of HMM states, K. K settings above 5 did not change the topographies of most common states. The corresponding results are shown in supplementary Figure S1. Once the HMM model was trained on the MEG data, we could obtain the state time courses and the spectral signature for each state. Figure 2 shows spatial power maps and temporal features of all states at the group level. Looking closely at the five mean activation maps averaged across 11 subjects, state 1 showed a large-scale activation in the frontal area in both hemispheres. States 2 and 3 corresponded to the left and right temporal activations respectively. State 4 corresponded to the parietal lobe and state 5 was expressed stronger in the occipital areas.

**Figure 2.**
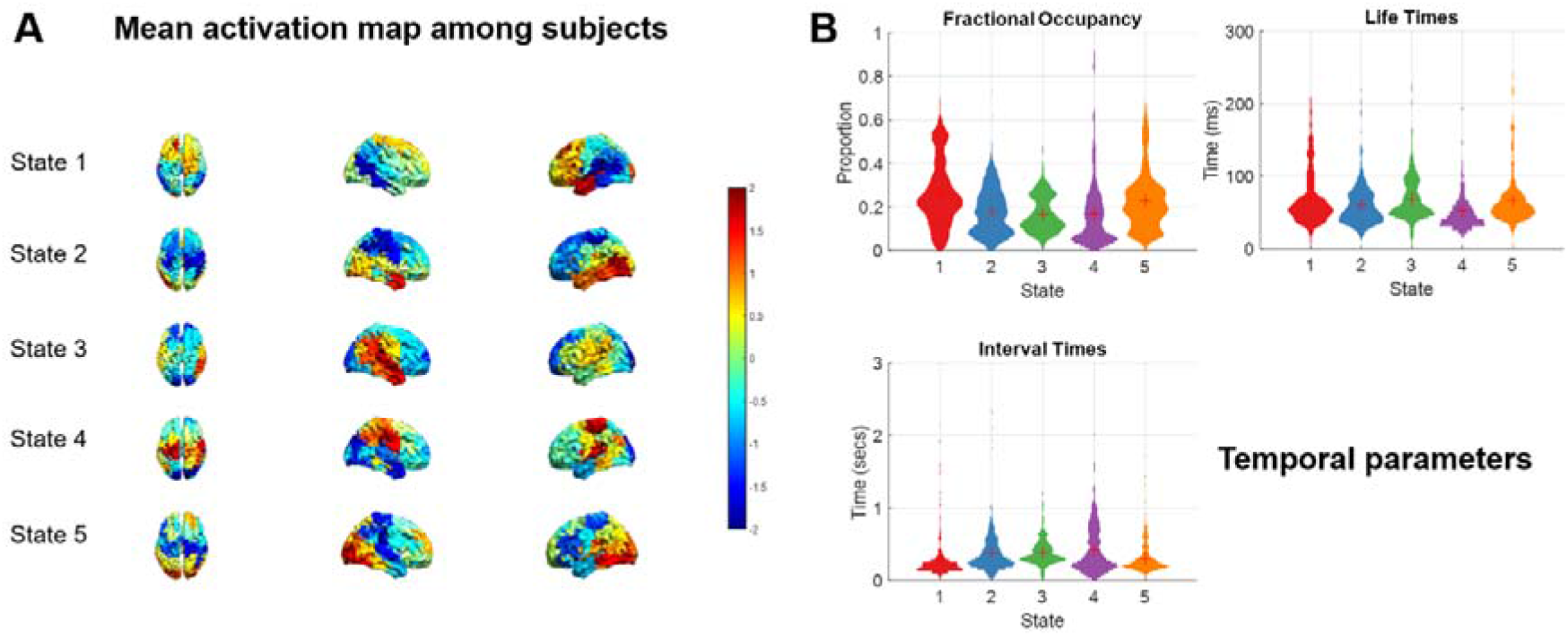
Outputs from running the HMM on the MEG data. A. Mean spectral activation maps of five states computed across all subjects in the 1-30Hz band. B. Distribution of temporal parameters of the five states: fractional occupancy, life times and interval times

Three parameters: fractional occupancy, life times and interval times were used here to illustrate the temporal statistics of each state. Fractional occupancy for a state is the proportion of time each subject spent in this state. The state life time refers to the number of time points per visit, known as the duration of visits to a state. This reflects the temporal stability of the states. The state interval time is the number of time points between subsequent visits. Five states had similar fractional occupancies around 15-25%. The state life times of all 5 states were around 50-100ms. The state interval times were also distributed similarly for the 5 states.

The state-wise correlation spectrum was computed for the time series of each iEEG bipolar channel based on the TDE-HMM results derived from MEG data. The correlation spectrum was defined as the correlation between the probability time course of a particular MEG-derived HMM state and the power envelope of the iEEG computed for each frequency bin (see Methods and Fig. 3A for details). The majority of iEEG channels had higher power correlations with the left and right temporal states (state 2 and state 3), which corresponded to the locations of contacts in the temporal clusters being much more common than elsewhere, as shown in Figure 3B. The spatial correspondence between HMM states and iEEG bipolar channel locations (Fig 3B) was determined by the smallest distance between the midpoint of the two iEEG contacts and the mean location of activated voxels for a particular state. Note that this calculation was done on individual rather than group topographies (see Supplementary Figure S2). Hence, the contact allocations do not always match the group state maps. As shown in Fig. 3A, the correlation between HMM probability time series of state 2 and 3 (from MEG data) and temporal spectral power (from iEEG data) was highest in theta/alpha band. Figure 4 shows spectrally resolved power maps of the two temporal states in different bands. States 2 and 3 were characterized by increased theta and alpha power in the left and right temporal areas respectively, consistent with the frequency range of highest correlations.

**Figure 3.**
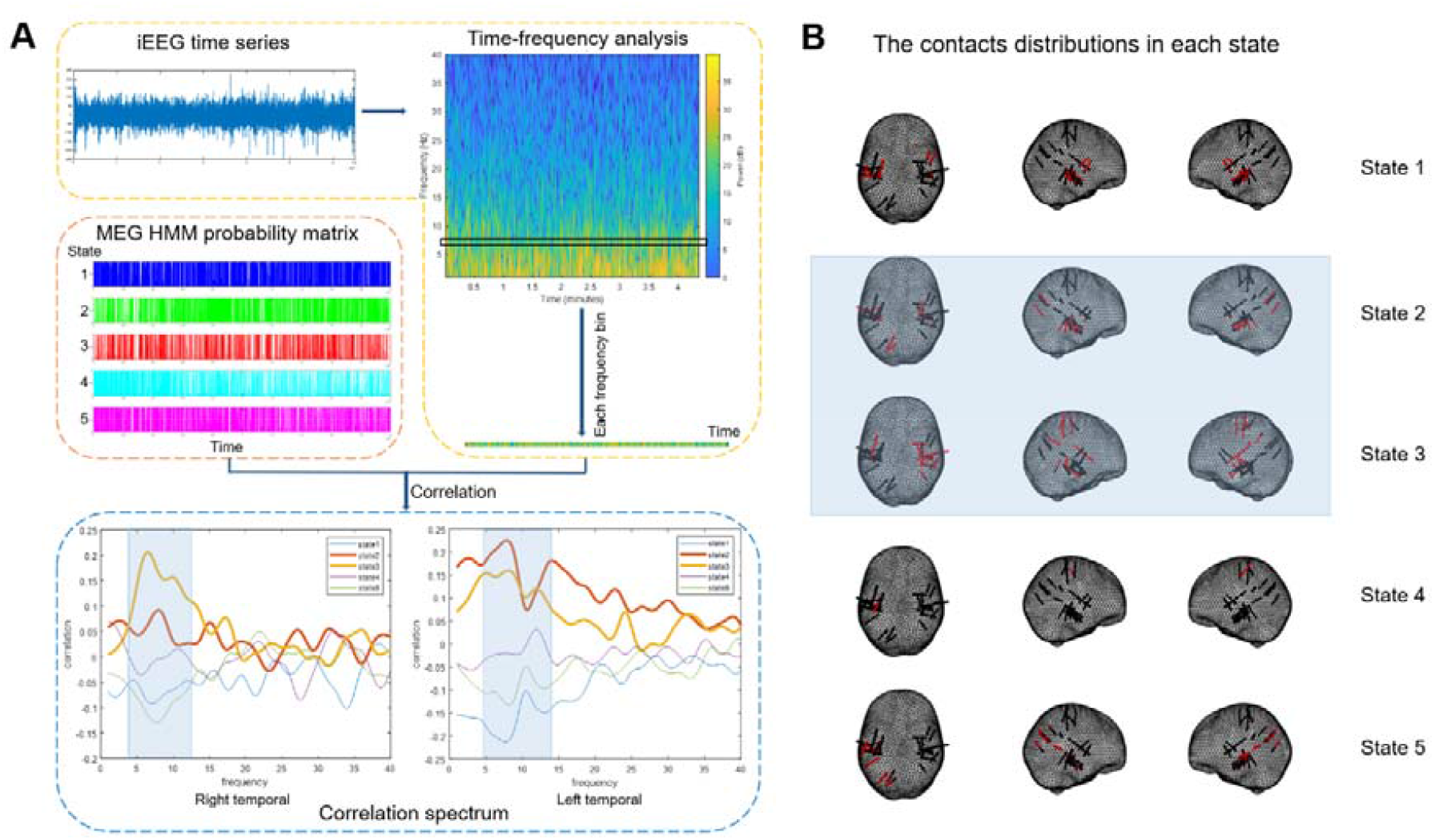
Combination of MEG-derived HMM results and iEEG data. A. The calculation of the cross-modality correlation spectrum, computed by correlating the MEG-derived HMM state time courses with the iEEG power time courses for all iEEG frequencies. B. The channel allocation to each MEG-derived HMM state according to the smallest distance.

**Figure 4.**
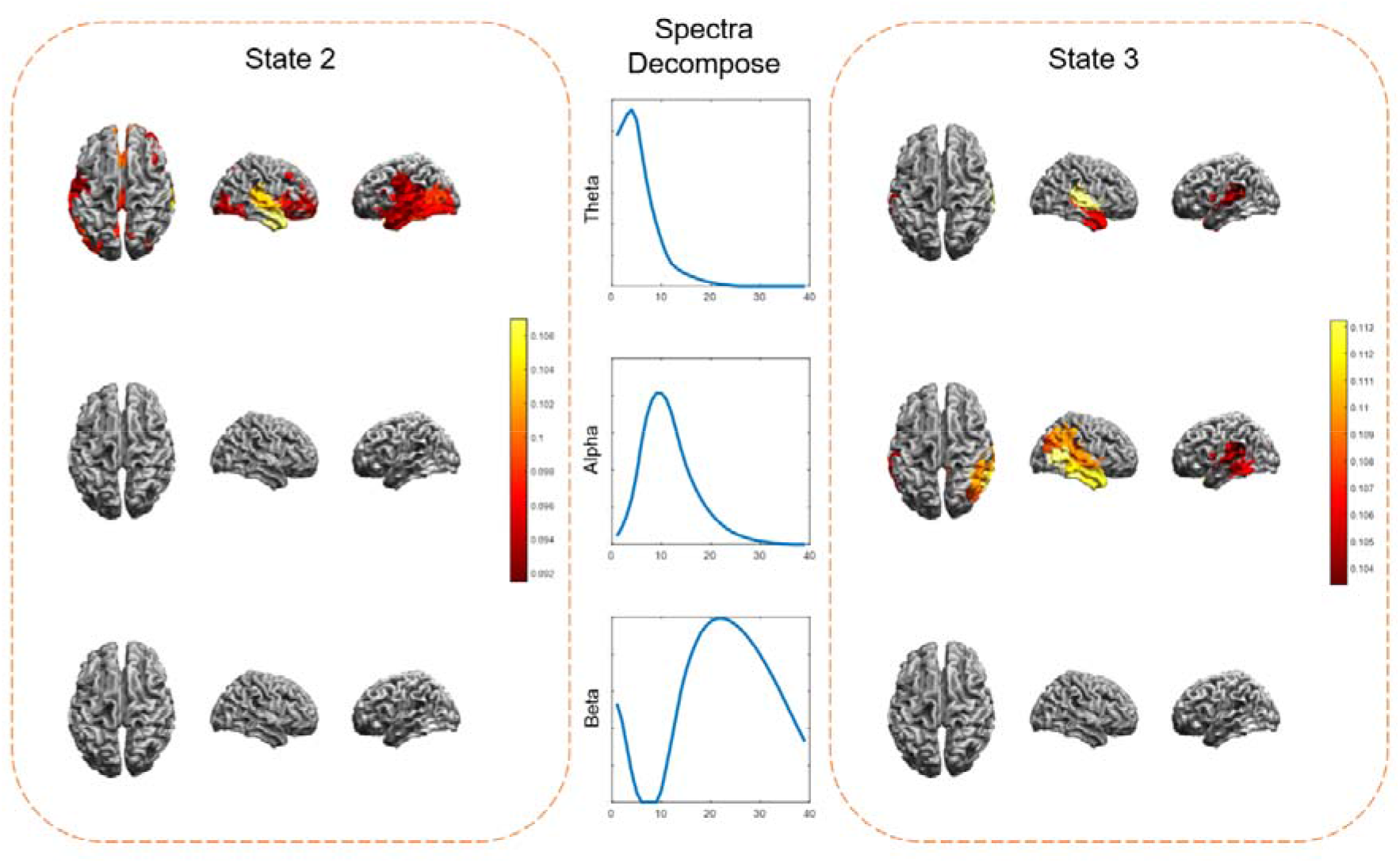
Frequency decomposition of two HMM inferred temporal states. The band limited power for state 2 and state3 showed that the majority of their power concentrated in the theta and alpha frequencies in the temporal cortex.

Finally, to make sure that the HMM computation was not driven by abnormal activity related to epilepsy, we repeated the analysis using HMM state topographies pre-computed from a set of healthy subjects (see Methods). The results are displayed in Figure 5. The activation maps looked very similar to those acquired from our HMM directly fit to patient MEG data as shown in Figure 5A. The similarity was quantified by Pearson correlation between the spatial activations of healthy fitted HMM output and our data-driven HMM output and was significant with *p*<0.01 for the corresponding states (Fig. 5B). This suggests that abnormal functional anatomy in patients does not preclude group HMM analysis.

**Figure 5.**
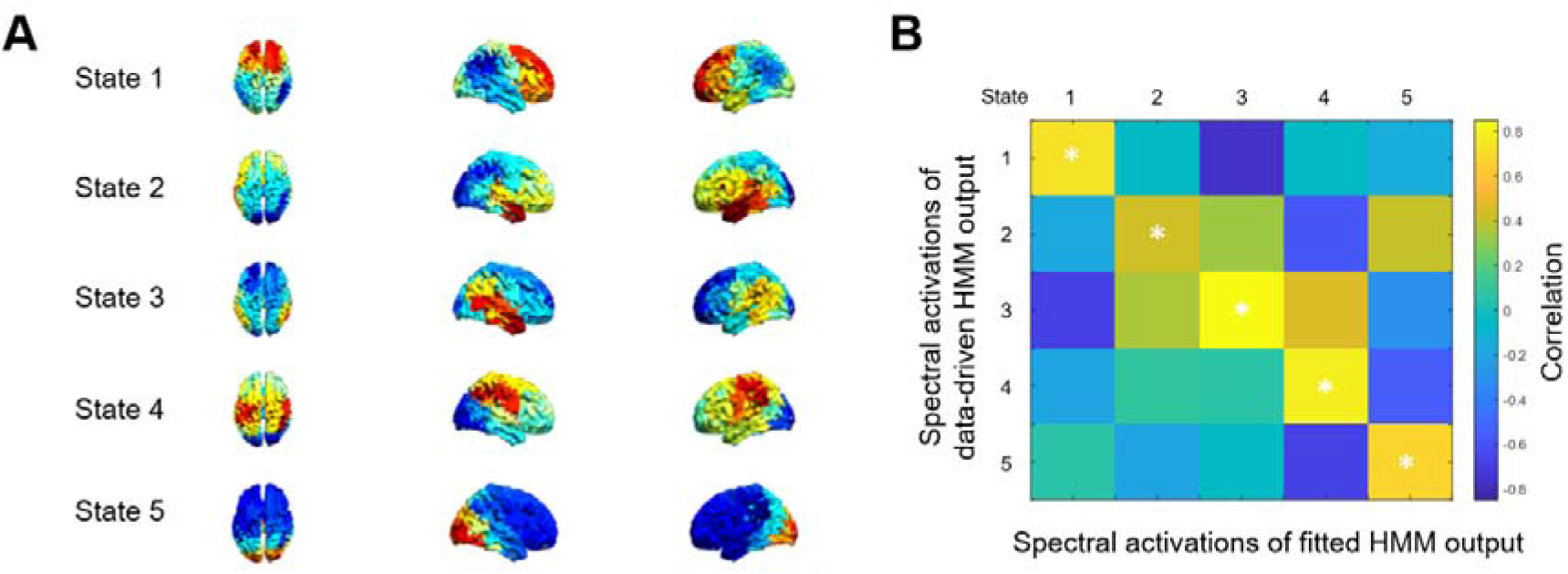
Validation of the patient MEG HMM inference using HMM inferred on a MEG dataset of healthy subjects. A. Mean spectral activation map of the epilepsy dataset estimated from healthy-cohort HMM fit. B. Pearson correlation coefficients between the spectral activations of healthy fitted HMM output and our data-driven HMM output.

## DISCUSSION

Non-invasive whole-brain MEG recordings could help put the iEEG data in the context of overall brain activity so that both modalities would maintain their inherent advantages whilst overcoming their limitations. By means of simultaneous MEG recording, large-scale networks in resting-state brain can be well described by repeated visits to short-lived transient brain states identified with TDE-HMM. This method provides information that is both spectrally and temporally resolved as different networks are described as being active or inactive at different points in time (Vidaurre et al., 2018). This is the first time HMM was applied to simultaneously recorded iEEG-MEG dataset and our pipeline (https://github.com/SiqiZhang0106/Dynamic-HMM-Analysis-on-Simultaneous-iEEG-MEG) could be used in future similar studies.

Five resting-state HMM states were inferred from the MEG dataset after removal of clearly abnormal interictal activity and exhibited evenly distributed temporal characteristics. Although iEEG is limited by inconsistent electrode coverage between patients, most of the implanted electrodes in our dataset were located in the temporal areas showing increased activity for states 2 and 3. The power time courses of iEEG electrodes were clearly related to HMM state time courses of the states whose spatial topography closely corresponded to the locations of the respective electrodes. The measure of concordance under simultaneous recordings constitutes a unique source of information on the coupling between depth and surface recordings (Dubarry et al., 2014).

The relative prevalence of temporal iEEG electrodes in our dataset is not surprising in light of the fact that temporal lobe epilepsy is the most frequent form of drug-resistant epilepsy referred to epilepsy surgery centres (Irena et al., 2017). EEG features in the theta/alpha band have been shown to differ between people with epilepsy and healthy controls (Yaakub et al., 2020). Our spectral analysis also showed that the two temporal states were mostly activated in the theta/alpha band. Each cortical area tended to preserve its own natural frequency, indicating that the observed oscillations reflect local physiological mechanisms (Rosanova et al., 2009). A number of observations made by means of intra-cerebral electrodes have illustrated the complexity of neuronal circuits that involve the temporal lobe (Kahane et al., 2015; Kahane and Bartolomei, 2010). Therefore, further research is necessary to clearly answer the question whether the activity driving these spectral correlations is healthy or epilepsy-related.

Our study is the first one to explore the dynamic spatiotemporal patterns in a simultaneous dataset focussing on presumably healthy rather than epileptic activity. It had several limitations, however. The sample size was limited because of the technical difficulty of acquiring simultaneous iEEG and MEG data although our dataset was quite large compared to other similar studies (Badier et al., 2017; Dalal et al., 2013). The locations of iEEG electrodes were variable among subjects which is typical of the iEEG setting. We only excluded data around clearly identified epileptic spikes which might have missed a large proportion of other pathological features. However, the comparison to HMM derived from a larger dataset of healthy subjects showed that this last limitation does not preclude HMM estimation from this kind of data.

## CONCLUSIONS

We showed for the first time that global brain states identified using HMM from non-invasive MEG data lending itself to group analysis have clear intracranial correlates. The correlation spectrum and concordance measures constitute important analysis pipeline and informational source on the relationship connecting depth and surface recordings. Although this fact is not surprising given that both MEG and iEEG data are generated by cortical activity, it forms the basis of further exploration of the possibly complex relations between the two kind of signals using HMM methods.

## Acknowledgements

The Wellcome Centre for Human Neuroimaging is supported by core funding from the Wellcome 203147/Z/16/Z. UK MEG community is supported by the MRC UKMEG Partnership grant MR/K005464/1. Qing Lu is funded by the Natural Science Foundation of China (81871066, 81571639); Jiangsu Provincial Medical Innovation Team of theProject of Invigorating Health Care through Science, Technology and Education (CXTDC2016004); Jiangsu Provincial key research and development program (BE2018609). Chunyan Cao is funded by Natural Science Foundation of China (81571346). Siqi Zhang is funded by China Scholarship Council (201806090144).

